# Validation of a deep learning model for automatic segmentation of skeletal muscle and adipose tissue on L3 abdominal CT images

**DOI:** 10.1101/2023.04.23.23288981

**Authors:** David P.J. van Dijk, Leroy F. Volmer, Ralph Brecheisen, Ross D. Dolan, Adam S. Bryce, David K. Chang, Donald C. McMillan, Jan H.M.B. Stoot, Malcolm A. West, Sander S. Rensen, Andre Dekker, Leonard Wee, Steven W.M. Olde Damink, Body Composition Collaborative

**Affiliations:** Department of Surgery, Maastricht University Medical Centre, Maastricht, The Netherlands; NUTRIM School of Nutrition and Translational Research in Metabolism, Maastricht University, The Netherlands; Department of Radiotherapy (MAASTRO), School of Oncology and Reproduction, Maastricht University, Maastricht, The Netherlands; Academic Unit of Surgery, School of Medicine, University of Glasgow, Glasgow Royal Infirmary, Glasgow, United Kingdom; Wolfson Wohl Cancer Research Centre, School of Cancer Sciences, University of Glasgow, Glasgow, United Kingdom; West of Scotland Pancreatic Unit, Glasgow Royal Infirmary, Glasgow, United Kingdom; Department of Surgery, Zuyderland Medical Centre, Sittard-Geleen, The Netherlands; Academic Unit of Cancer Sciences, Faculty of Medicine, University of Southampton, Southampton, UK; Department of General, Visceral and Transplant Surgery, University Hospital Aachen, Aachen, Germany; Department of Nutrition and Movement Sciences, School of Nutrition and Translational Research in Metabolism (NUTRIM), Faculty of Health, Medicine and Life Sciences, Maastricht University, Maastricht, The Netherlands; Department of Obstetrics and Gynaecology, Maastricht University Medical Center, Maastricht, The Netherlands; Department of Medical Oncology, University of Southampton, Southampton, UK; Department of Surgery, University of Southampton, Southampton, UK; UCL Cancer Institute, University College London, London, UK; Department of General Surgery, Portsmouth Hospitals University NHS Trust, Portsmouth, UK

**Keywords:** body composition, deep learning, convolutional neural networks, image segmentation, cancer cachexia, computed tomography

## Abstract

**Background:** Body composition assessment using abdominal computed tomography (CT) images is increasingly applied in clinical and translational research. Manual segmentation of body compartments on L3 CT images is time-consuming and requires significant expertise. Robust high-throughput automated segmentation is key to assess large patient cohorts and ultimately, to support implementation into routine clinical practice. By training a deep learning neural network (DLNN) with several large trial cohorts and performing external validation on a large independent cohort, we aim to demonstrate the robust performance of our automatic body composition segmentation tool for future use in patients.

**Methods:** L3 CT images and expert-drawn segmentations of skeletal muscle, visceral adipose tissue, and subcutaneous adipose tissue of patients undergoing abdominal surgery were pooled (n = 3,187) to train a DLNN. The trained DLNN was then externally validated in a cohort with L3 CT images of patients with abdominal cancer (n = 2,535). Geometric agreement between automatic and manual segmentations was evaluated by computing two-dimensional Dice Similarity (DS). Agreement between manual and automatic annotations were quantitatively evaluated in the test set using Lin’s Concordance Correlation Coefficient (CCC) and Bland-Altman’s Limits of Agreement (LoA).

**Results:** The DLNN showed rapid improvement within the first 10,000 training steps and stopped improving after 38,000 steps. There was a strong concordance between automatic and manual segmentations with median DS for skeletal muscle, visceral adipose tissue, and subcutaneous adipose tissue of 0.97 (interquartile range, IQR: 0.95-0.98), 0.98 (IQR: 0.95-0.98), and 0.95 (IQR: 0.92-0.97), respectively. Concordance correlations were excellent: skeletal muscle 0.964 (0.959-0.968), visceral adipose tissue 0.998 (0.998-0.998), and subcutaneous adipose tissue 0.992 (0.991-0.993). Bland-Altman metrics (relative to approximate median values in parentheses) indicated only small and clinically insignificant systematic offsets : 0.23 HU (0.5%), 1.26 cm^2^.m^-2^ (2.8%), -1.02 cm^2^.m^-2^ (1.7%), and 3.24 cm^2^.m^-2^ (4.6%) for skeletal muscle average radiodensity, skeletal muscle index, visceral adipose tissue index, and subcutaneous adipose tissue index, respectively. Assuming the decision thresholds by Martin et al. for sarcopenia and low muscle radiation attenuation, results for sensitivity (0.99 and 0.98 respectively), specificity (0.87 and 0.98 respectively), and overall accuracy (0.93) were all excellent.

**Conclusion:** We developed and validated a deep learning model for automated analysis of body composition of patients with cancer. Due to the design of the DLNN, it can be easily implemented in various clinical infrastructures and used by other research groups to assess cancer patient cohorts or develop new models in other fields.

## Introduction

Body composition assessment using routine abdominal computed tomography (CT) images is increasingly applied in clinical and translational research. By measuring the tissue area at the level of the third lumbar vertebra (L3) and scaling for subject height, precise assessments of total body mass of skeletal muscle (SM), visceral adipose tissue (VAT), and subcutaneous adipose tissue (SAT) can be made.^1^ Body composition has been found to be highly independently predictive of survival, especially among cancer patients. In particular, low skeletal muscle mass (i.e., sarcopenia), low adipose tissue mass, and decreased skeletal muscle radiodensity (i.e., myosteatosis) have been shown to be associated with shorter overall survival in various cancer types.^2–4^

Body composition exhibits substantial heterogeneity among people due to natural variation in age, sex, race, and build.^5^ These intrinsic inter-personal differences are unrelated to disease and may therefore obscure disease related body composition effects, necessitating large population-based data cohorts to adjust for them.

Manual segmentation of body compartments on L3 CT images is time-consuming and requires significant expertise. Therefore, robust high-throughput automated segmentation is key to body composition assessment in large patient cohorts and ultimately, to support implementation of body compositon assessment into routine clinical practice. A deep learning neural network (DLNN) can be an essential part of such an automated workflow.

One challenge for developing a robust DLNN is that patients do not always have the ideal CT scans for body composition assessment, such that variable orientation of the patient, degradation of image quality due to radiation artefacts, and individual-specific anatomical attributes may result in poor performance of an automated segmentation algorithm.^6^ A systematic review revealed that one in three DLNN studies of body composition segmentation have been developed with less than 100 unique human subjects, and more than half of the reviewed studies used exclusively single-institutional datasets.^7^ Robust DLNNs need to be trained on datasets that are large enough to incorporate the heterogeneity created by a variety of scanners, image acquisition settings, image reconstruction kernels, patient positioning protocols, and sufficiently high heterogeneity of subject clinical presentations. Additionally, the quantitative performance of DLNNs need to be comprehensively evaluated with external test datasets sourced from a wholly independent clinical workflow and a separate clinical setting from the one used to train the DLNN.^8^

In previous work, the DLNN that is the subject of this paper had been independently validated using a large polytrauma patient cohort extracted from the same university hospital, albeit at a different department and for a clinically distinct setting.^9^ This was nonetheless considered a challenging validation attempt due to the large variation in patient positioning (including arms and hands appearing inside the field of view) as well as radiation artifacts (e.g., from metal devices attached to the patient). Even with this challenging cohort, the present DLNN model performed very well.

A robust, fully inter-institutional and large-scale external testing with useen datasets is needed for developing a quality AI tool for potential clinical use. This paper presents the first validation of the Mosamatic DLNN in a surgical oncology cohort using data from a separate hospital, using previously unseen scanners, with independent radiology scan protocols, and with reference delineations provided by independent clinicians.

## Patients and methods

### Patients

**A total of 3,187 patients requiring abdominal surgery who had undergone a CT scan prior to surgery contributed by 32 distinct centres were used for DLNN development (see general patient characteristics in Table 1).** These comprised of de-identified data abstracted from previously ethics board-approved clinical studies; permission for secondary analysis was obtained via the principal investigators of the respective studies. We used L3 CT slices from: three colorectal liver metastases trials - two from multiple sites across the UK and a single-institution study in The Netherlands; two ovarian cancer trials among five participating Dutch centers; and one pancreatic cancer trial of patients operated either in Aachen, Germany, or in Maastricht, the Netherlands.

**Table 1.**
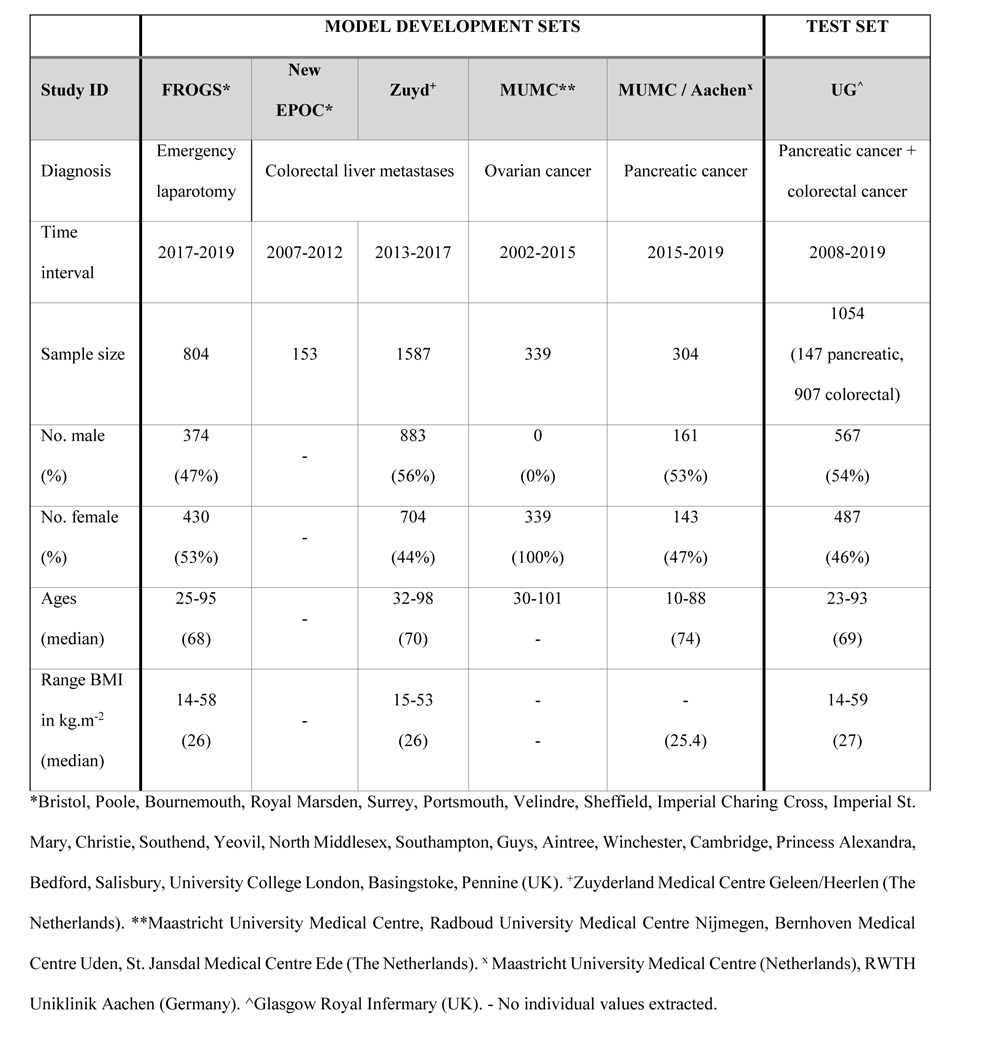
General patient characteristics for the deep learning neural network development sets and the external test set.

|  | MODEL DEVELOPMENT SETS |  |  |  |  | TEST SET |
| --- | --- | --- | --- | --- | --- | --- |
| Study ID | FROGS* | New EPOC* | Zuyd<sup>+</sup> | MUMC** | MUMC / Aachen<sup>x</sup> | UG<sup>^</sup> |
| Diagnosis | Emergency laparotomy | Colorectal liver metastases |  | Ovarian cancer | Pancreatic cancer | Pancreatic cancer + colorectal cancer |
| Time interval | 2017-2019 | 2007-2012 | 2013-2017 | 2002-2015 | 2015-2019 | 2008-2019 |
| Sample size | 804 | 153 | 1587 | 339 | 304 | 1054<br>(147 pancreatic, 907 colorectal) |
| No. male (%) | 374<br>(47%) | - | 883<br>(56%) | 0<br>(0%) | 161<br>(53%) | 567<br>(54%) |
| No. female (%) | 430<br>(53%) | - | 704<br>(44%) | 339<br>(100%) | 143<br>(47%) | 487<br>(46%) |
| Ages (median) | 25-95<br>(68) | - | 32-98<br>(70) | 30-101<br>- | 10-88<br>(74) | 23-93<br>(69) |
| Range BMI in kg.m<sup>-2</sup> (median) | 14-58<br>(26) | - | 15-53<br>(26) | -<br>- | -<br>(25.4) | 14-59<br>(27) |

\*Bristol, Poole, Bournemouth, Royal Marsden, Surrey, Portsmouth, Velindre, Sheffield, Imperial Charing Cross, Imperial St. Mary, Christie, Southend, Yeovil, North Middlesex, Southampton, Guys, Aintree, Winchester, Cambridge, Princess Alexandra, Bedford, Salisbury, University College London, Basingstoke, Pennine (UK). <sup>+</sup>Zuyderland Medical Centre Geleen/Heerlen (The Netherlands). \*\*Maastricht University Medical Centre, Radboud University Medical Centre Nijmegen, Bernhoven Medical Centre Uden, St. Jansdal Medical Centre Ede (The Netherlands). <sup>x</sup> Maastricht University Medical Centre (Netherlands), RWTH Uniklinik Aachen (Germany). <sup>^</sup>Glasgow Royal Infirmary (UK). - No individual values extracted.

An independent external validation set comprised 2,535 L3 CT slices at different time intervals taken from 1,054 unique subjects diagnosed with either resectable colorectal or pancreatic cancer (see Table 1).^10,11^ Ethical approval was granted by the West of Scotland Research Ethics Committee, Glasgow.

### Image acquisition and reference segmentations

The aforementioned datasets comprised CT scans from a broad range of equipment vendors and image acquisition settings. Images were archived in DICOM (Digital Imaging and Communications in Medicine) format. Table S1 (see online supplementary materials) summarizes the diverse imaging settings as recorded in DICOM metadata.

All human-made segmentations in this study were created with *Slice-o-matic* (Tomovision, Quebec, Canada). Regions of interest (ROIs) were defined using standardized Hounsfield Unit (HU) ranges (SM: - 29 to +150, VAT: -150 to -50, SAT: -190 to -30). Absolute areas were normalized by physical height squared to derive skeletal muscle index (SMI), visceral adipose tissue index (VATI), and subcutaneous adipose tissue index (SATI). Mean HU in SM at L3 was used as the skeletal muscle radiation attenuation (SMRA). All human reference segmentations were made by clinical researchers trained to perform body compostion analysis in Slice-o-matic.

Previously published analyses on the external validation dataset had been made with ImageJ (National Institutes of Health, v1.47, http://rsbweb.nih.gov/ij/), but this method was shown to overestimate adipose tissue areas relative to other software.^12^ Every validation subject in this study was therefore independently re-annotated in Slice-o-matic by the original data owners. To ensure consistency for direct comparison, we re-computed areas and mean HU for all subjects with independent Python code, and confirmed equivalent values with each version of Slice-o-matic used to 2 decimal places or better.

### Deep learning neural network (DLNN)

A DLNN for multi-label segmentation of SM, VAT, and SAT was built from a canonical 2D U-Net,^13^ with minor change in the input layer to match the dimensions of a CT slice (512x512). An essential development for this work was to chain two independently-trained U-Net networks; the first U-Net was developed to segment the whole abdomen, whilst ignoring hands, arms, CT mattress and extraneous medical devices that sometimes appeared in the CT field of view. The second U-Net was specialized for segmenting SM, VAT, and SAT within the abdominal outline detected by the first U-Net (see online supplementary materials Figure S1 and its accompanying text).

Pixel intensities were clipped to the range [-500, +500] HU for the abdomen segmentation network. The reference abdominal region was generated by computing the outermost continuous contour of the human expert’s SAT region before morphologically filling in every pixel inside. The range of intensities was further clipped to [-200, +200] HU to train the multi-label segmentation of muscle and fat. In each network, clipped intensities were scaled between [0,1] via standard min-max normalization. Pre-processed CT images where stored and handled in DICOM format. Human expert segmentations were extracted from Slice-o-matic in its proprietary TAG format and converted to Python (NumPy) array objects before training the deep learning model.

Hands, arms, and other extraneous objects were rare within the training set, thus we synthetically over-sampled images with extraneous objects outside the abdomen until they comprised 50% of each training batch while developing the abdomen U-Net. To train the muscle and fat multi-level segmentation network, all available 3,187 subjects were randomly shuffled and split into 80% for training and 20% for validation. Given the relatively large sample size, a (non-overlapping) 80-20 split is superior to alternative methods like K-fold cross-validation where each validation block ultimately ends up being “seen” by the training algorithm, potentially introducing bias due to data leakage. More details of DLNN construction have been provided in online supplementary materials.

CT slices and human-drawn (reference) annotations for the external validation were not revealed until the final DLNN model had been selected and all its model weights permanently fixed. Pre-processing of the test set followed the same steps as aforementioned. The full DLNN code (stripped of all trained models and patient data) is made open access (see data availability statement). **The trained algorithm can run easily on a conventional office laptop with standard specifications.**

### Automatic L3-selection

For use on large cohorts and for ease of future clinical implementation, automatic vertebra localization is necessary. We have integrated a state-of-the-art externally validated and open-source tool known as TotalSegmentator (https://github.com/wasserth/TotalSegmentator).^14,15^ In keeping with the “narrow AI” paradigm, we have chained together highly specialized AI tools for each task. TotalSegmentator was first used for automated segmentation of all visible vertebrae in a volumetric CT study. The resulting labelled masks were used to locate all the slices intersecting L3, and then we selected the CT slices closest to the centre of the segmented object (see Figure S2).

### Analysis

Geometric agreement was evaluated by using 2D Dice Similarity (DS) comparing the DLNN segmentations of SM, SAT, and VAT against the corresponding annotation made by human experts. DS computes the area of the intersection between human and DLNN segmentations as a fraction of half the summated area (human-drawn area plus DLNN-drawn area). Perfect geometric agreement implies DS = 1, and if the intersection area is zero then DS = 0. Agreement of SMI, VATI, SATI, and SMRA between manual and automatic annotations were quantitatively evaluated in the test set using Lin’s Concordance Correlation Coefficient (CCC) and Bland-Altman’s Limits of Agreement (LoA) (with and without repeated measurements). By using the human-drawn annotations in the test set as reference and then applying the risk classification supplied by Martin et al,^2^ we computed the diagnostic performance (sensitivity, specificity, balanced accuracy, and agreement kappa) of the DLNN results. Statistical analyses were performed in R (version 4.2.0).

## Results

### Model training

Total loss and DS curves in the training dataset show DLNN model convergence within about 40,000 steps (see Figure S3). There was rapid improvement within the first 10,000 steps but DS was largely stable thereafter. Total (Dice+L2) loss continued to decrease gradually but we stopped model training after 38,000 steps, since there was very little to gain with further training. The DLNN weights after the last training step were thus fixed as the “final model” for subsequent testing. The established segmentation tool was named MosaMatic.

### Segmentation speed

The DLNN was able to segment a single CT-image in around 2 seconds and the whole external validation cohort in around 90 minutes. Considering that an experienced clinical researcher trained in body composition analysis needs a minimum of 2-5 minutes to segment a single CT-image, the use of automatic segmentation can potentially save months of work when assessing large cohorts.

### Concordance between manual and DLNN segmentations

The overall distribution of DS for SM, VAT, and SAT in the quarantined validation dataset are summarized in the box-whisker plot shown in Figure 1(a). The median DS for SM was 0.97 (interquartile range, IQR: 0.95-0.98), with a tail of outliers down to a minimum DS of 0.45. The distributions of DS for VAT (median: 0.98, IQR: 0.95-0.98) and SAT (median: 0.95, IQR: 0.92-0.97) were highly skewed, with extreme outliers landing near zero (these were patients with very small amounts of total adipose tissue). The DS is known to be overly sensitive for small volumes, and this can also be seen in our results – Figure 1(b, c, and d). Lin’s CCC evaluation of SMRA, SMI, VATI, and SATI comparing expert segmentations (as reference) and DLNN results (as test) was excellent, as shown in Figure 2 (a-d). Numerical measures of the concordance correlation coefficient (CCC), bias correction factor for slope of agreement, and finally the

**Figure 1:**
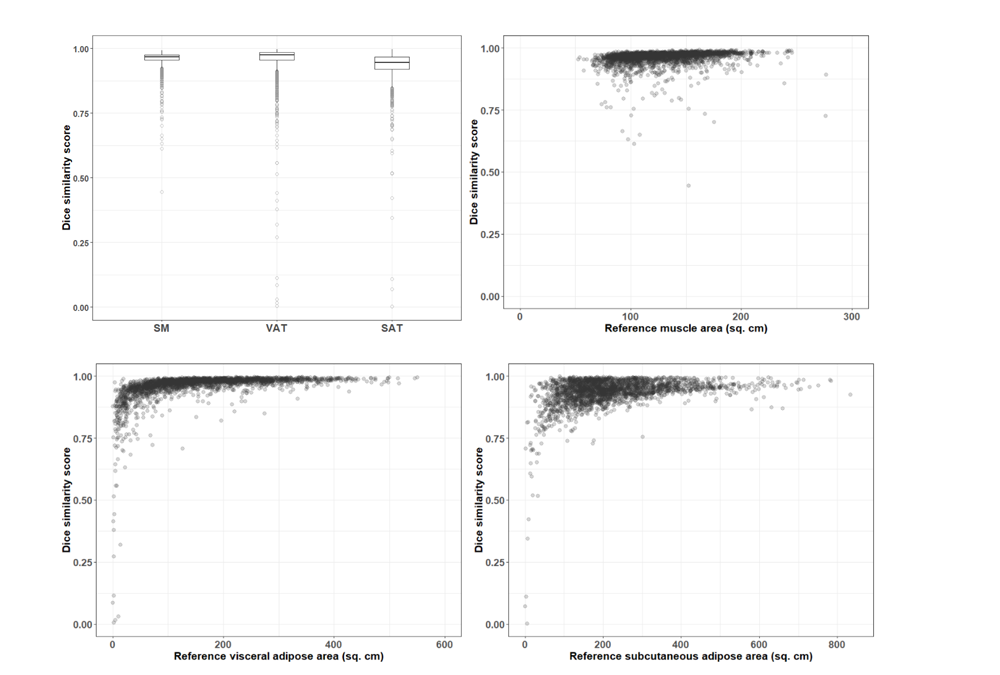
Distribution of geometric DS on L3 slice for skeletal muscle (SM), subcutaneous fat (SAT), and visceral fat (VAT) (a) Box-whisker plot showing the median DS as the solid horizontal line and the interquartile range as the upper and lower limits of the box. The vertical line ends indicate 1%-tile and 99%-tile, and outliers outside this range are shown as individual dots. (b) – (d) show the distribution of DS as a function of SM area, VAT area, and SAT area, respectively.

**Figure 2:**
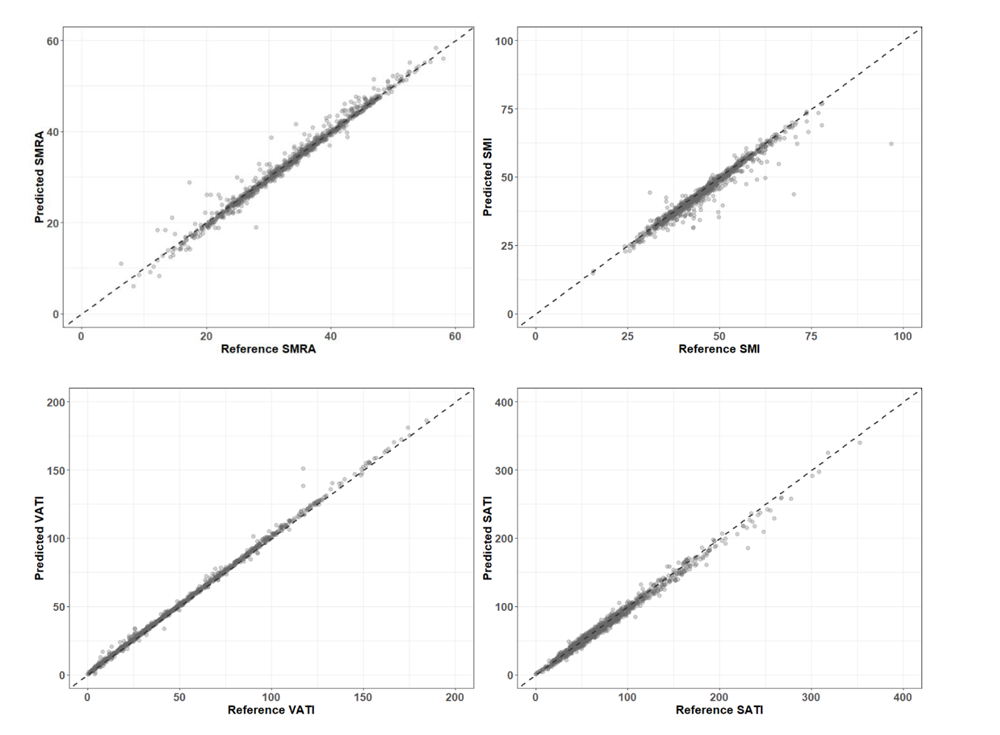
Lin’s concordance correlation (CCC) plots. (a) skeletal muscle attenuation (SMRA), (b) skeletal muscle index (SMI), (c) visceral fat index (VATI) and (d) subcutaeous fat index (SATI). The units of SMRA are HU. The units of SMI, VATI, and SATI are all cm^2^.m^-2^. Reference values were defined as those extracted from human-drawn segmentations. Predicted values were extracted from DLNN-made segmentations.

Bland-Altman intervals of agreement without repeated scans are provided in Table 2. The CCC ranges from 0.964 (for SMI) up to 0.998 (for VATI). The errors in the agreement slope, as indicated by deviation from the dotted line in Figure 2, were all close to unity, indicating no major deviations from the ideal, which is supported by bias correction multipliers being better than 0.991 (i.e. no correction implies 1.00). Based on our large cohort, median *in vivo* values (which are in reality age-and sex-dependent) of SMRA, SMI, VATI and SATI roughly fall in the vicinity of 50 HU, 45 cm^2^.m^-2^, 60 cm^2^.m^-2^ and 70 cm^2^.m^-2^. The Bland-Altman metrics (with percentages in parentheses) indicate only small systematic offsets of 0.23 HU (1.0%), 1.26 cm^2^.m^-2^ (2.9%), 1.02 cm^2^.m^-2^ (2.5%), and 3.24 cm^2^.m^-2^ (4.9%) for SMRA, SMI, VATI, and SATI, respectively. The upper and lower limits of the Bland-Altman tests indicate SATI had the widest random variation component (-6.7 to 13 cm^2^.m^-2^). Most importantly for risk stratification by muscle fat content, the random noise component of SMRA was estimated at about 2 to 3 HU in magnitude, and correspondingly for SMI about 3 to 5 cm^2^.m^-2^ in magnitude.

**Table 2.** Concordance correlation, bias correction factor, and Bland-Altman agreement without repeated measures (n = 1054)

| Bland-Altman estimates of agreement for primary scan only (n = 1054) |  |  |  |
| --- | --- | --- | --- |
|  | Concordance correlation<br>(95% confidence interval) | Bias correction<br>factor | Bland-Altman agreement<br>(95% lower-upper limits) |
| SMRA | 0.991 (0.990 – 0.992) | 0.999 | 0.23 (-2.06 – 2.52) HU |
| SMI | 0.964 (0.959 – 0.968) | 0.991 | 1.26 (-3.11 – 5.63) cm<sup>2</sup>·m<sup>-2</sup> |
| VATI | 0.998 (0.998 – 0.998) | 0.999 | -1.02 (-4.55 – 2.50) cm<sup>2</sup>·m<sup>-2</sup> |
| SATI | 0.992 (0.991 – 0.993) | 0.997 | 3.24 (-6.69 – 13.2) cm<sup>2</sup>·m<sup>-2</sup> |
| Bland-Altman estimates of agreement for repeated scans only (n = 449) |  |  |  |
|  | Concordance correlation<br>(95% confidence interval) | Bias correction<br>factor | Bland-Altman agreement<br>(95% lower-upper limits) |
| SMRA | 0.991 (0.990 – 0.992) | 0.999 | 0.18 (-2.08 – 2.45) HU |
| SMI | 0.973 (0.969 – 0.976) | 0.997 | 0.75 (-3.56 – 5.06) cm<sup>2</sup>·m<sup>-2</sup> |
| VATI | 0.998 (0.998 – 0.998) | 0.999 | -1.07 (-4.55 – 2.41) cm<sup>2</sup>·m<sup>-2</sup> |
| SATI | 0.992 (0.991 – 0.993) | 0.998 | 2.55 (-8.36 – 13.4) cm<sup>2</sup>·m<sup>-2</sup> |

### Consistent concordance for repeated measurements

In 449 subjects, we obtained a repeated CT image at varying time intervals ranging from within a month up to 12 months. Whereas the scope of this study was not to objectively quantify longitudinal precision, we can already derive some preliminary insight into stability with repeated imaging over time using this data. The concordance plots for SMRA, SMI, VATI, and SATI for *repeated scans* are equivalent to Figure 2 (see Figure S4). There was no evidence of divergence from the high concordance observed in the agreement on primary CTs. According to CCC metrics and Bland-Altman limits with repeated measures, there are no notable changes between agreement of body composition indices between primary (top half of Table 2) and repeat scans (bottom half of Table 2).

### Accuracy

We tested the clinical significance of using the DLNN segmentations with respect to a change in stratification for sarcopenia and low SMRA using the widely used thresholds defined by Martin et al.^2^

Overall accuracy of stratification was 0.93 for sarcopenia (sensitivity: 0.99, specificity: 0.87) and 0.98 for low SMRA (sensitivity: 0.98, specificity: 0.98). The discretized agreement (Cohen’s inter-rater kappa) was 0.85 for sarcopenia and 0.96 for low SMRA, which is generally considered as being excellent. For completeness, a 2x2 confusion matrix for sarcopenia and low SMRA is included in the online supplemental materials as Figure S5.

### Automatic L3-selection

We tested the accuracy of L3 mid-slice localization from TotalSegmentator using a small independent test cohort of 30 subjects. The tool correctly extracted the CT-slice at L3 in 30 out of 30 cases (100%).

## Discussion

In this study, we present our high performing and externally validated deep learning model for automated segmentation of CT-based L3 slices. Due to its excellent performance in both internal and external validation cohorts, the DLNN-generated segmentation can reliably replace manual segmentation when performing body composition assessment. This opens up new possibilities both in clinical and scientific settings, such as cost-and time-effective clinical implementation and large cohort/population studies. Clinically and subject to clinical implementation study to follow this work, our automated L3 body composition segmentation tool is intended to be easily implemented in standard practice for all routine CT-scans, which clinicians can then use for prognostic risk assessment and treatment decision making. Changes in body composition over time can be detected during oncologic follow-up, which might provide early indications of treatment effect or disease progression/recurrence. Going from a prognostic tool to a predictive tool – in which the tool is used for treatment decisions - still remains a large step to take as large international data-sets are needed to provide clinical reference values.

Body composition is highly variable among sex, age, race, and cancer types.^3,4,16–18^ For this reason, developed clinical cut-offs vary greatly among different patient cohorts and prognostic models of outcome (e.g. survival) are likely to fail during external validation.^3,19^ In addition, body composition can be dependent on other clinical parameters and may have stronger prognostic effects when combined with parameters such as systemic inflammation and weight loss.^10,20,21^ We have previously demonstrated that such combinations or “host phenotypes” are more predictive of overall survival than tumor-based prognostic scores in patients with colorectal liver metastases.^20^ **Larger cohorts are needed for each cancer type, as these could support the use of body composition analysis in the standard diagnostic work-up, and potentially aid in clinical treatment decision-making.** Automated body composition analysis is the only way of acquiring sufficient data for adequate Z-scoring and accounting for the aforementioned patient characteristics. While cut-offs are necessary for clinical use, we advocate the development of a clinical risk calculator, as the prognostic effect of body composition variables are incremental^4^ and should therefore not be arbitrarily forced into dichotomic cut-offs. **In the end, integrating body composition data with established prognostic factors such as tumour stage may improve prediction of a patient’s prognosis. A combined tumour and host focused approach would provide a basis for clinical trials aimed at exploring whether body composition-based prognostic information can be used as a basis for treatment decision making (e.g. palliative intent instead of curative intent, or indication for/selection of (neo)adjuvant therapy).**

Scientifically, our L3 segmentation tool enables assessment of large (incl. historical) cohorts that would be unfeasible to segment manually. In addition, as the AI has learned from multiple observers, it has not learned an expert’s specific signature, ensuring a more stable output. However, the true value of automated segmentation is that it facilitates the inclusion of body composition as a study parameter in RCTs, as the time and effort of analysis is reduced from a couple of months to a few minutes. This enables stratification and selection of patients with different body compositions, creating either homogenous or heterogeneous cohorts as required. Including body composition is particularly important in oncology as it is related to chemotherapy effectiveness and toxicity.^22^ Ideally, chemotherapy dosing should be based on lean mass to prevent dose-limiting toxicities for which DLNN would be a logical application in the future.

Some other automated segmentation tools have been developed. The largest cohort (n=12,128) was used for development of the AI tool published by Magudia et al.^16^ Their tool performed well with similar dice scores to our algorithm. Their training cohort only included 604 pancreatic cancer patients while the large (n= 12,128) hospital dataset was used to derive reference curves. However, the large hospital dataset only included patients without cancer and cardiovascular disease, making it less applicable to a clinical population of subjects with cancer who frequently display body composition alterations. In addition, analysis of CT-scans of cancer patients can be more challenging due to anatomic abnormalities and suboptimal patient positioning. As patients with cancer were excluded, the tool by Magudia et al. could perform worse in cancer cohorts. **Our analyses did not exclude patients with anatomical variations or unconventional patient positioning, which prevents overfitting the model to a specific patient group and will likely result in a more robust segmentation tool**. Dabiri et al. published an automated segmentation tool which was trained on two cohorts of patients with cancer (n=2529).^23^ Their segmentation tool performed similarly well compared with our segmentation tool. However, in contrast to our study, they did not perform external validation, making it uncertain how their AI performs in other cohorts.

For volumetric CTs as input, an important consideration is how to select the slice intersecting the middle of the L3 vertebra, and more generally in case the user arbitrarily wishes to select some other vertebra. In keeping with the “narrow AI” paradigm, we have elected to implement a modular software design such that highly specialized DLNN are joined up sequentially in a workflow to accomplish a meaningful task. For the present, we integrated the state-of-the-art and validated TotalSegmentator tool to automatically localize spinal vertebrae. If a superior vertebrae segmentation tool should emerge in future, we could relatively easily adapt our workflow to incorporate the new tool, compared to “all-in-one” monolithic software design. **While the DLNN showed excellent performance, even with challenging CT-scans, it has its limitations. In particular, analysis of CT-scans of patients with anatomical abnormalities (e.g. large abdominal hernia, colostomy, profound edema) or of patients with abnormal/non-standard positioning in the CT-scanner can lead to (partially) incorrect segmentations. Such challenging CT-images should then be manually corrected and stored prospectively. In due time, this cohort of** “challenging CT-images” can be used to retrain and improve the DLNN. In addition, different deep learning segmentation algorithms will have different limitations depending on the cohort. A comparative study using both healthy individuals and different patient groups could provide insight into how these different algorithms perform and if one algorithm is preferred over the other in specific cohorts.

The key step forward will be implementing automated segmentation into clinical practice and making it easily accessible for new research initiatives. Our tool was created in such a way that it can be easily integrated in clinical imaging software or work independent alongside existing imaging infrastructure. To ensure easy access for research purposes, the untrained AI will be freely available for scientific use and the trained AI can be used under license through a web-app or docker by other research groups. This enables rapid implementation and much needed data collection to develop clinical prediction tools.

## Conclusion

In this study, we developed a reliable deep learning model that was externally validated for automated analysis of body composition of patients with cancer. **To simplify future use and potential integration of the DLNN-based automated segmentation workflow, we have incorporated the steps into a web browser-based graphical user interface. Clinical implementation within our own institution is not within the scope of this study, but is the subject of a future study. For external institutional users who wish to access the trained model for research, please see data availability statement.**

## Supplemental material

Please see the attached document for online access.

## Data availability statement

This work concerns only secondary re-use of clinical study data of patients, which were obtained in deidentified form with permission from the original principal investigators. Each study had previously been reviewed by a competent ethics body. Data may be obtained from the aforementioned principal investigators upon reasonable request. Source code for data preparation of CT slices and human reference annotations, along with the DLNN model architecture, are publicly available here under a Creative Commons 4.0 CC-BY-NC License: https://github.com/MaastrichtU-CDS/BodyCompL3_DLNN_Open_Code

The trained model is available upon request through www.mosamatic.com.

## Data Availability

All data produced in the present study are available upon reasonable request to the authors.

## Acknowledgments

The New EPOC study was supported by Cancer Research UK.

