## Supplemental_Materials for "Validation of a deep learning model for automatic segmentation of skeletal muscle and adipose tissue on L3 abdominal CT images" (from another version of this preprint: version 2, 2023-08-01)

**Supplementary (Online) Materials**

**Table S1. Image acquisition settings in model development and test sets, as gleaned from DICOM metadata.**

|  | MODEL DEVELOPMENT SETS | | | | | | TEST SET |
| --- | --- | --- | --- | --- | --- | --- | --- |
| Study ID | **FROGS*** | **New EPOC*** | **Zuyd^+^** | | **MUMC**** | **MUMC / Aachen^x^** | **UG^^^** |
| Diagnosis | Emergency laparotomy | Colorectal liver metastases | | Ovarian cancer | | Pancreatic cancer | Pancreatic cancer + colorectal cancer |
| Vendor(s) | GE  Siemens  Toshiba | GE  Philips  Siemens Toshiba | GE  Philips  Siemens  Toshiba | | Siemens | - | GE  Philips  Siemens  Toshiba |
| Energy (kVp) | 80-140 | 100-140 | 80-140 | | 80-140 | 100-130 | 80 - 140 |
| Current (mA) | 80-1078 | 67-696 | 47-645 | | 50-640 | 50-625 | 41 - 739 |
| Pixel area (mm^2^) | 0.31-0.97 | 0.33-1.87 | 0.24-1.87 | | 0.30-1.87 | 0.31-1.87 | 0.29-1.87 |
| Slice thickness (mm) | 0.6-5.0 | 0.6-8.0 | 1.0-8.0 | | 2.0-10.0 | 2.0-10.0 | 0.5 - 10.0 |

* Bristol, Poole, Bournemouth, Royal Marsden, Surrey, Portsmouth, Velindre, Sheffield, Imperial Charing Cross, Imperial St. Mary, Christie, Southend, Yeovil, North Middlesex, Southampton, Guys, Aintree, Winchester, Cambridge, Princess Alexandra, Bedford, Salisbury, University College London, Basingstoke, Pennine (UK). ^+^ Zuyderland (The Netherlands). ** Maastricht, Radboud Nijmegen, Bernhoven, St. Jansdal, Ede (The Netherlands). ^x^ Maastricht University Medical Centre (Netherlands) and RWTH Uniklinik Aachen (Germany). ^ University Glasgow Medical Centre (UK). - No individual values extracted from DICOM scan metadata. Slice thickness implies reconstructed and interpolated slice width; all scanners used were multi-slice helical CT.

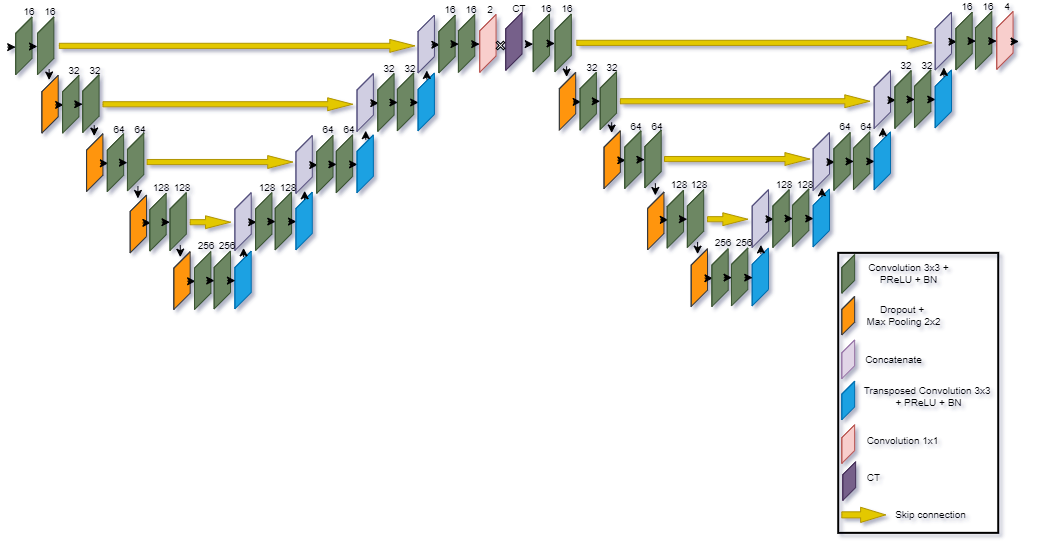

**Figure S1. A schematic representation of our pipeline with both U-Net structures for abdominal outline on the left side chained with the fat/muscle multilevel body component segmentation on the right side.**

Our architecture closely follows a canonical U-Net architecture, but with a few optimizations for this specific task. An essential development for this work was to chain two independently-trained U-Net networks; the first U-Net was developed to segment the whole abdomen, whilst ignoring hands, arms and extraneous objects that does sometimes appear in the CT field of view. The second U-Net was specialized for segmenting SM, SAT and VAT within the abdominal outline detected by the former. The U-Net has been so-named due to its symmetrical structure of stepwise image compression into a bottleneck layer of artificial neurons, followed by an expanding pathway until its output has the same dimensions as the input. The essential function of the compressive pathway is to down-sample the image to lower resolution for computational efficiency with higher degree of feature abstraction. Blocks in the downsampling path each consisted of two convolution layers with pixel padding of 1 so as to maintain consistent size of input to the next layer. The expanding pathway comprised transposed convolutions concatenated with earlier levels of abstracted information via “skip” connections. Parametric Rectified Linear Units (PReLU) were used as activation functions everywhere except the final convolution layer and we initialized our weights using the He initialization method. The final The final convolution layer was followed by softmax activation to extract probability density maps for four segmentation labels (SM, VAT, SAT or other). Batch normalization was applied after every convolutional layer and L2 (i.e. ridge) regularization was added to reduce the effects of overfitting. Dropout layers were inserted after every block of convolutions to further reduce the chance of overfitting. Identical training hyperparameters were applied in both U-Nets. Adam was used as an optimizer to train the network with parameters β1 = 0.9, β2 = 0.000 and ε = 10^-8 which correspond to standard default values. L2-regularization is set at 0.0001 and dropout at 0.05. We trained our model to a maximum of 100000 steps with learning rate fixed at 0.0001. Early stopping was engaged if no improvement were made in 10000 steps.

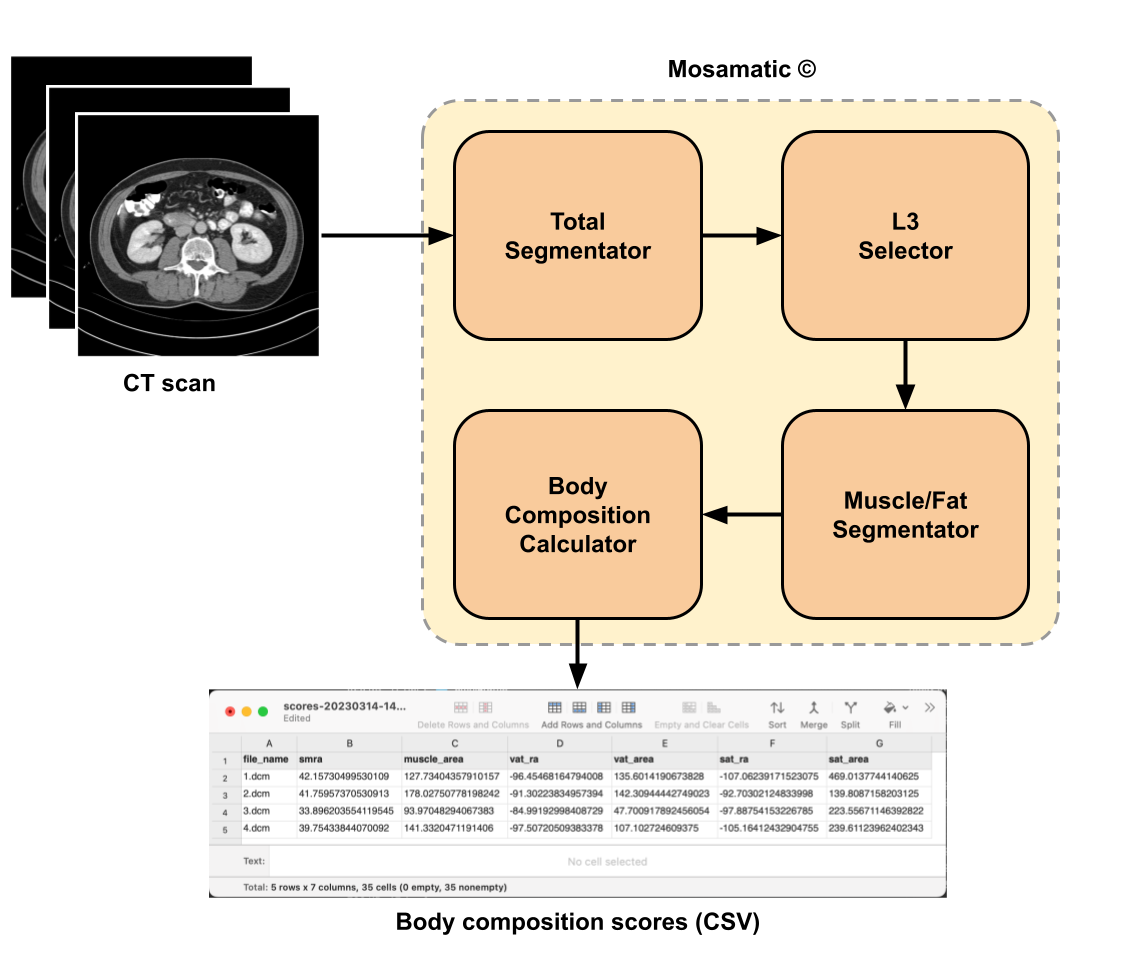

**Figure S2. Integration of automatic L3 selection**

As input, the tool needs to be provided with a set of abdominal CT-scans. As output, the tool produces a comma-separated value (CSV) file containing the body composition scores for each CT-scan. Inside the tool, we use a sequence of steps starting with the open-source tool Total Segmentator (used as a Python package) that extracts 104 anatomical structures from the CT-scan including the individual spinal vertebrae of which the L3 Selector identifies the L3 vertebra and selects the DICOM slice intersecting the middle of the L3 vertebra. The Muscle/Fat Segmentator subsequently extracts the muscle and fat regions from the image, after which the Body Composition Calculator calculates the body composition scores, as mentioned previously, and outputs the CSV table.

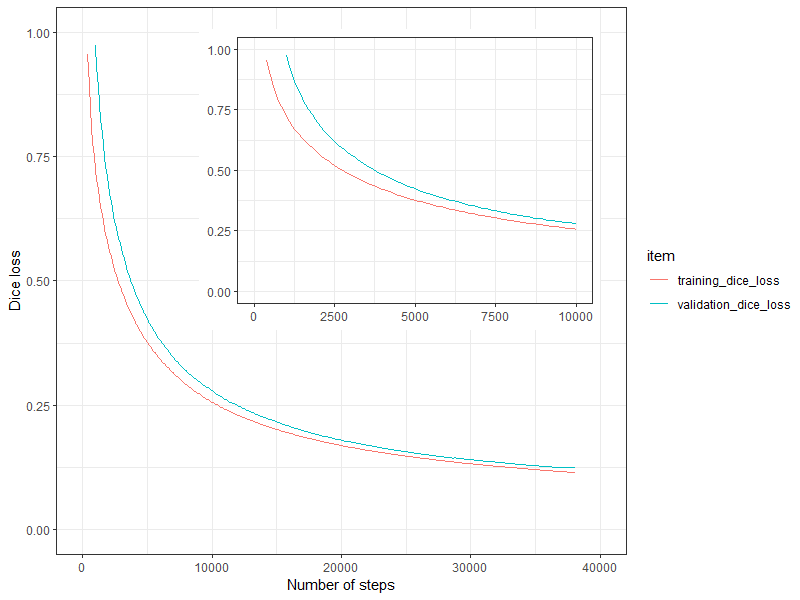

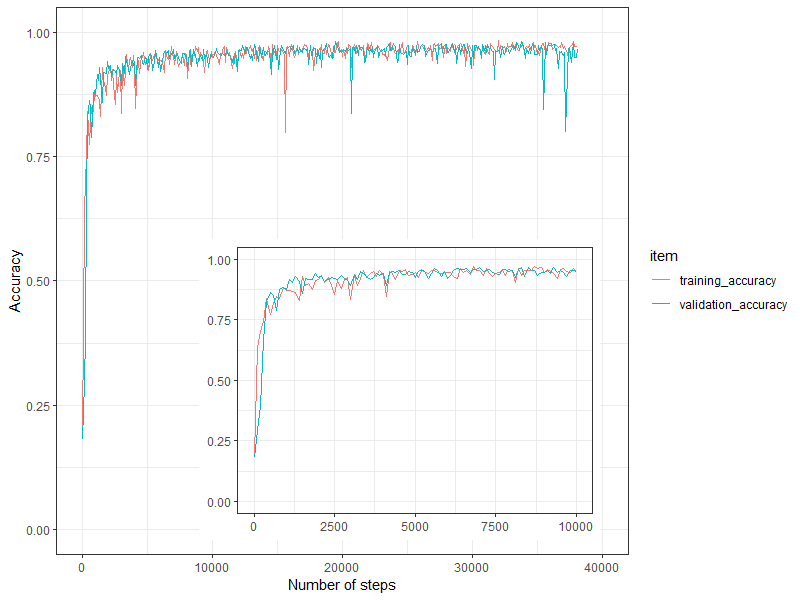

**Figure S3. Training and validation loss curves.**

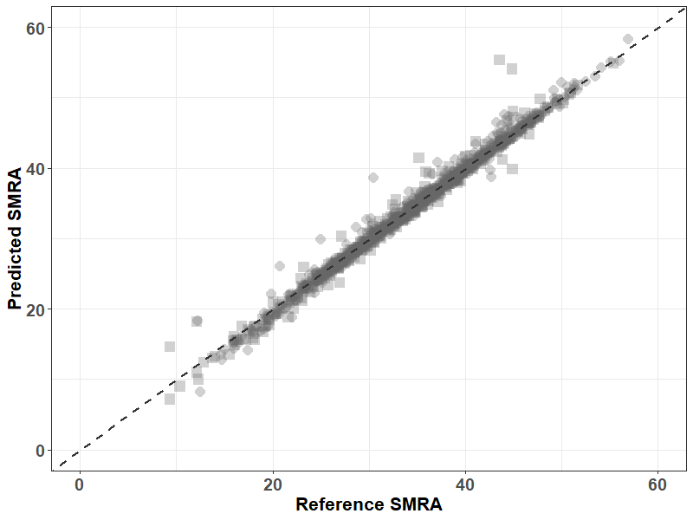

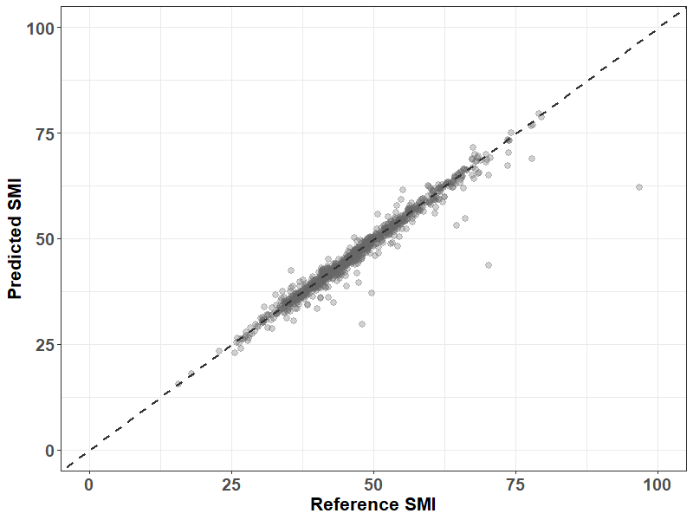

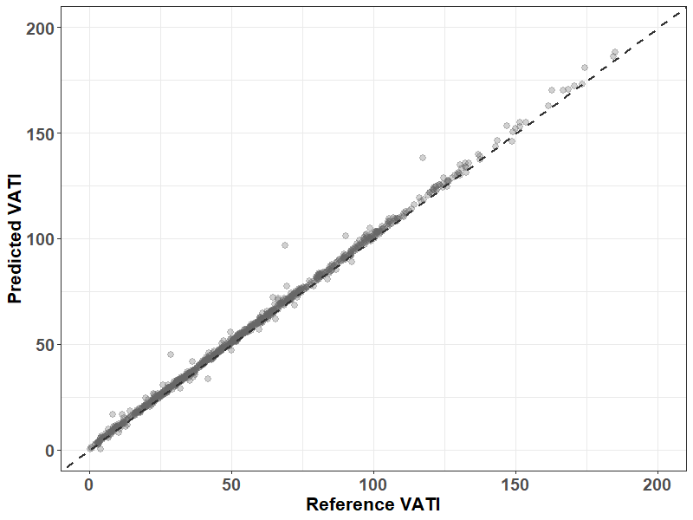

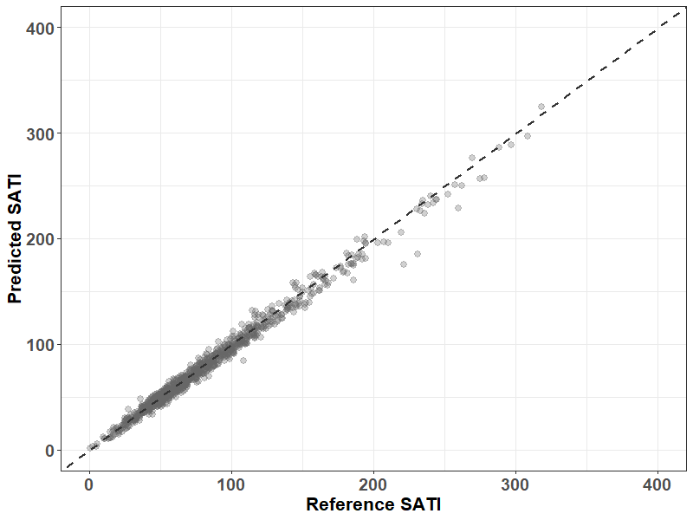

**Figure S4: Box-and-Whisker plot of the DSC on L3 slice for muscle, subcutaneous fat and visceral fat**

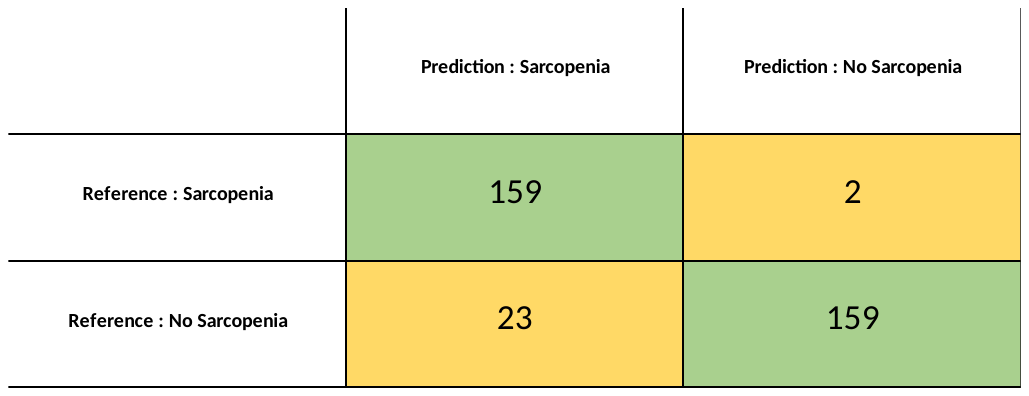

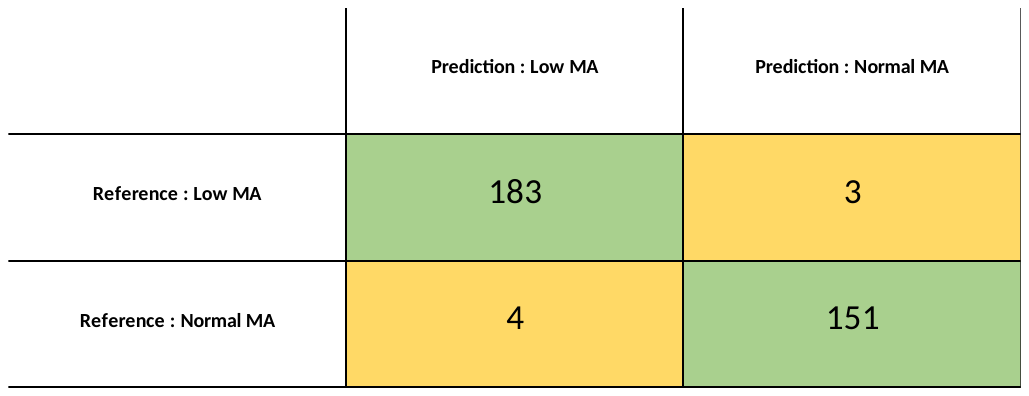

**Figure S5: Confusion matrices for classification of Sarcopenia and Low Muscle Attenuation, assuming the thresholds proposed by Martin et al.**
